# Refining reproduction number estimates to account for unobserved generations of infection in emerging epidemics

**DOI:** 10.1101/2021.11.08.21266033

**Authors:** Andrea Brizzi, Megan O’Driscoll, Ilaria Dorigatti

## Abstract

**Background:** Estimating the transmissibility of infectious diseases is key to inform situational awareness and for response planning. Several methods tend to overestimate the basic (R_0_) and effective (R_t_) reproduction numbers during the initial phases of an epidemic. The reasons driving the observed bias are unknown. In this work we explore the impact of incomplete observations and underreporting of the first generations of infections during the initial epidemic phase.

**Methods:** We propose a debiasing procedure which utilises a linear exponential growth model to infer unobserved initial generations of infections and apply it to EpiEstim. We assess the performance of our adjustment using simulated data, considering different levels of transmissibility and reporting rates. We also apply the proposed correction to SARS-CoV-2 incidence data reported in Italy, Sweden, the United Kingdom and the United States of America.

**Results:** In all simulation scenarios, our adjustment outperforms the original EpiEstim method. The proposed correction reduces the systematic bias and the quantification of uncertainty is more precise, as better coverage of the true R_0_ values is achieved with tighter credible intervals. When applied to real world data, the proposed adjustment produces basic reproduction number estimates which closely match the estimates obtained in other studies while making use of a minimal amount of data.

**Conclusions:** The proposed adjustment refines the reproduction number estimates obtained with the current EpiEstim implementation by producing improved, more precise estimates earlier than with the original method. This has relevant public health implications.

**Summary:** We propose a back-imputation procedure tackling the issue of unobserved initial generations of infections to reduce the bias observed in the early *R*_*0*_ and *R*_*t*_ estimates and apply it to EpiEstim using simulated and reported COVID-19 data to evaluate it.

## Introduction

The wide-ranging impacts of the current COVID-19 pandemic have highlighted the threats posed by infectious diseases to our society. The impacts of such threats are not exclusively confined to the domain of public health: in addition to the millions of confirmed deaths worldwide, COVID-19 has caused severe economic and societal disruption around the world. As such, it is crucial that the properties of all emerging pathogens are adequately characterised as soon as possible, making the best use of the limited data that are typically available in the early phases of an epidemic. In particular, understanding the transmissibility of a novel pathogen allows for the evaluation of the risks involved, to inform public health decision making and the implementation of interventions. In this context, the basic and instantaneous reproduction numbers, *R*_*0*_ and *R*_*t*_, represent key epidemiological parameters. *R*_*t*_ represents the average number of secondary infections caused by a single infectious individual at time *t* and *R*_*0*_ represents the average number of secondary infections generated by a typical infection in a completely susceptible population. These parameters relate to important quantities such as the final size of an epidemic [1] and the critical herd immunity threshold [2,3] and are essential to project the expected future number of cases, hospitalisations and deaths. In the last couple of decades, several methods have been developed to estimate R_t_. EpiEstim, developed by Cori et al. [4], has been recommended as one of the best methods for near real-time estimation of *R*_*t*_ to detect changes in transmissibility patterns [5]. However, EpiEstim and other commonly used statistical methods, can suffer from systematic overestimation of the basic reproduction number in the early stages of an epidemic [6]. In the initial epidemic stages, EpiEstim is outperformed by simpler inference methods based on exponential growth [7], which produce smaller bias and better quantify uncertainty in *R*_*0*_ estimates. Following the theory of exponential growth [7], we propose an adjustment to EpiEstim to account for missing initial generations of infections and use simulated data to test its effectiveness. We show how the adjustment entails a reduction in the bias of both the *R*_*0*_ and early *R*_*t*_ estimates produced by the original EpiEstim method. Finally, we compare the estimates produced with and without the proposed adjustment on early COVID-19 data in Europe and in the US.

## Methods

Several methods, based upon different frameworks, have been developed to estimate the reproduction number. One of the simplest methods assumes exponential growth of the number of new infections, which can be characterised by applying a simple linear regression on the log-transformed incidence data. Wallinga and Teunis [8] base their methodology on determining likelihoods of chains of infections, White and Pagano [9] use branching process theory, while Bettencourt and Ribeiro [10] base their approach on SIR differential equations. In this work we focus on EpiEstim, the method developed by Cori et al [4], which has been recommended for near real-time estimation [5] and has been applied extensively during the ongoing and past epidemics [11].

Despite differences in the mathematical formulations, all methods described above infer *R*_*t*_ or *R*_*0*_ from case incidence data using assumptions on the generation interval or the serial interval distributions. The generation interval is defined as the average length of time between the moment an individual becomes infected and the moment in which they infect a secondary case. Similarly, the serial interval corresponds to the difference between symptom onsets of the primary case and symptom onset of the secondary case. Assuming that infectiousness starts after symptom onset and the infectiousness profile of an individual is independent of the incubation period, the two distributions are equivalent [4]. Therefore, without loss of generality, we focus on the generation interval distribution, denoted *w*.

### Exponential Growth

One of the simplest methods to characterise the speed of an epidemic, as measured by the growth rate *r*, is to fit the log-transformed incidence data by linear regression. The growth rate *r* can then be used to estimate the reproduction number *R*_*0*_ if the generation interval *w* is known [12]. When observations with 0 cases are present, linear regression requires the addition of a small constant to each incidence data point, so that the logarithm of each data point is well defined. The assumption of exponential growth is justifiable in a first epidemic period where the proportion of susceptible individuals in a population is large. As more individuals get infected and immunity accumulates in the population, the growth of the number of new cases slows down and deviates from being exponential. A rule of thumb to identify the time window of the exponential growth, proposed by White and Pagano [9] and based on the theoretical work of Ball and Donnelly [13], is that the cumulative number of infected individuals does not exceed the square root of the population size.

### EpiEstim

Cori et al. [4] model observed infections as a Poisson process, where the mean is defined via two quantities: the effective reproduction number *R*_*t*_, and population infectiousness 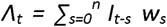. The reproduction number is assumed to remain constant over sliding windows of time with length *τ*, allowing for data aggregation over time and reduced variance. Assuming a Gamma prior distribution on *R*_*t,τ*_, the posterior distribution is Gamma distributed with parameters:

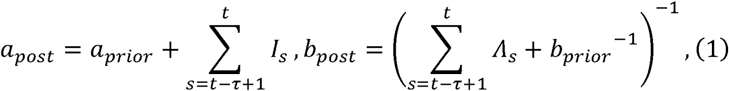

where *a* and *b* refer to the shape and scale parameters, respectively. Considering *τ* = 1 and uninformative priors (i.e. letting *a*_*prior*_, *b*_*prior*_ tend to 0) yields the posterior mean:

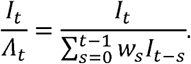

Thus we can effectively think of EpiEstim’s estimates of *R*_*t*_ as closely related to the ratio between the number of observed infections on day *t* over the population infectiousness on the same day. Given enough data, this estimator can accurately identify *R*_*t*_ and detect abrupt changes in transmissibility [5]. However, the method can suffer from systematic bias in the initial period of estimation, as described by O’Driscoll et al. [6]. When the first chains of infections are not observed, the estimator will tend to attribute all new cases to the first observed cases, overestimating the reproduction number.

### EpiEstim - proposed adjustment

We propose an adjustment to account for unobserved initial infections and exponential growth similar to the one developed in Dorigatti et al. [14]. Specifically, we assume that the epidemic is growing exponentially to back-impute infections in the period prior to the fist observation. Note that this assumption is theoretically justified in the early stages of an epidemic [7], and exponential growth inference methods are between the most accurate in this initial period, as observed in O’Driscoll et al.[6]. Our procedure can be summarised in three simple steps and is visualised in Figure 1. First, we fit a linear model on the log-transformed incidence data to estimate the growth rate,, as shown in Figure 1A. Second, we use this linear model to back impute incidence data, prior to the time of the first observed case. In particular, we obtain an estimate of the number of cases for *S* days, where *S* is the largest possible generation interval length, i.e., the largest value such that *w*_*s*_ > 0. We highlight that it is not necessary to round the output of the inferred number of cases, and estimates lower than 1 should not be removed. Finally, we apply EpiEstim to the extended epidemic curve (including the back imputed incidence data).

**Figure 1.**
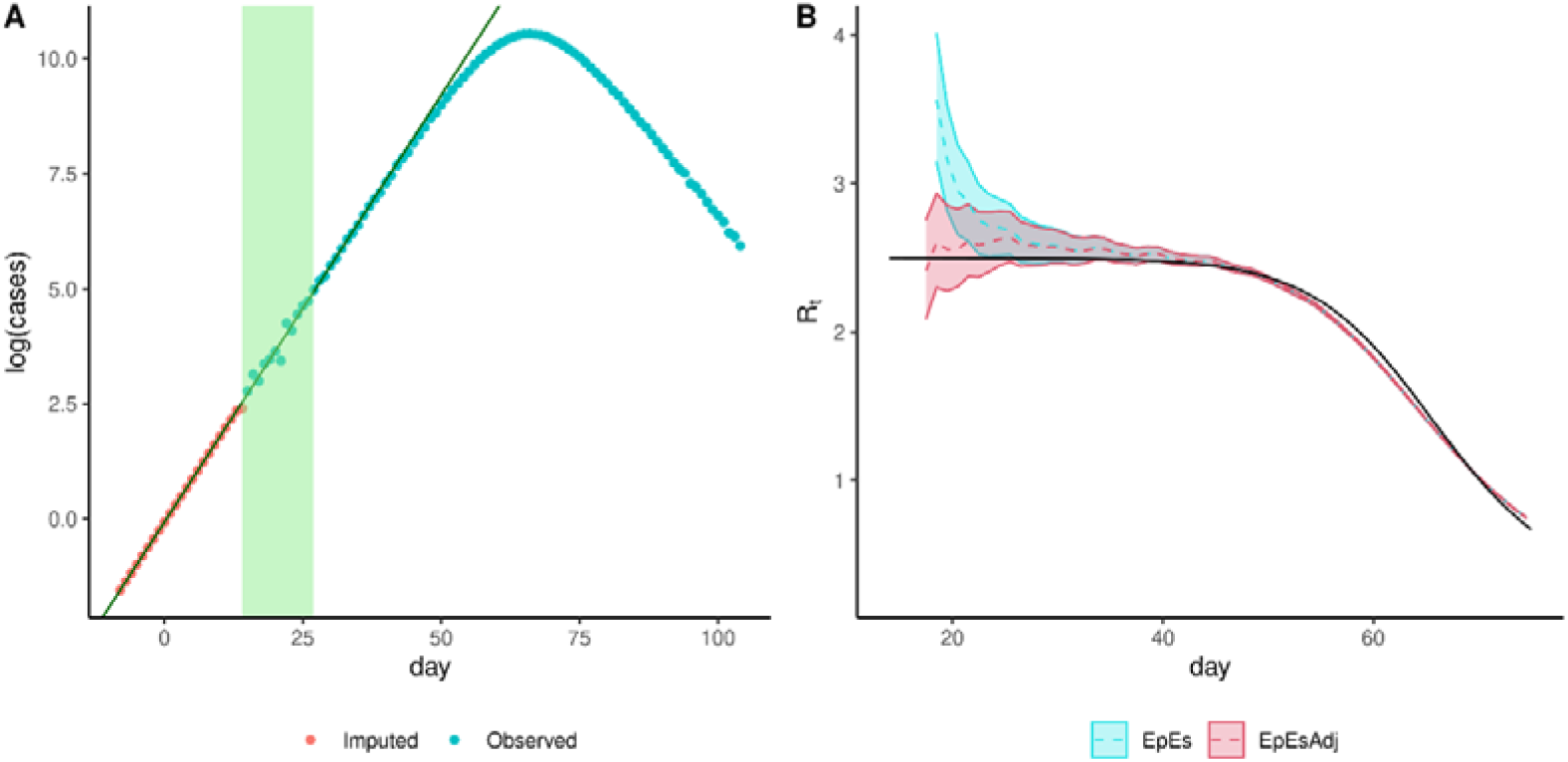
Visualisation of the exponential growth adjustment method applied to simulated data obtained with R_0_ = 2.5, reporting rate ρ = 1 and resulting changes in posterior estimates having assumed 2 unobserved generations. On panel A, the logarithms of the first reported data (those in the green region) are used to fit a linear model (line). The linear model is then used to back-impute unobserved cases (red dots) to complement the available data (blue dots). On panel B, the true R_t_ value (black solid line) is compared to EpiEstim estimates without (blue) and with (red) adjustment using sliding windows of 7 days. The back-imputation reduces the initial mean estimates (dotted lines) and 95% credible interval widths (ribbons). The adjusted method then converges to the original method as the importance of the imputed datapoints vanishes. Method abbreviations: EpiEstim (EpEs); Adjusted EpiEstim (EpEsAdj)

### Simulations

To evaluate the effects of our correction, we compared the performance of EpiEstim with and without the adjustment on simulated data. We additionally compared the results to those obtained by fitting a linear exponential growth model, as the performance of our correction strongly depends on the accuracy of the estimates of the growth rate. We used a stochastic SEIR simulator to generate 100 epidemic curves for each *R*_*0*_ values in {1.5, 2,2.5,3}.

We considered a large population of 10^6^ individuals, and epidemics initiated by 5 initial infections. We assumed 3 days and 3.5 days as the mean times spent in the exposed and infectious compartments respectively, yielding a 6.5 mean generation interval commonly used to model COVID-19 [6].

To account for imperfect reporting, we simulated two types of issues commonly affecting the observed data: underreporting and unobserved initial generations of infections. We simulate unobserved initial generations by considering the simulated data from day 15 to day 28, corresponding to two generation intervals worth of data after having missed the first two generations.

Further, we simulated under-reporting by considering a constant reporting rate ρ in {0.15, 0.3, 0.5, 1}. Daily observations were sampled as binomial realisations of true incidence with success probability equal to the reporting rate ρ.

## Results

### Method comparison

We fitted the exponential growth, EpiEstim and adjusted EpiEstim methods to the simulated data assuming a Gamma distributed generation interval matching the mean and variance of the generating process (i.e., 6.5 mean generation interval). Figure 2 shows the effect of our adjustment when the reporting rate is 50% and shows that the initial bias observed in the estimates obtained with EpiEstim is strongly reduced by the proposed adjustment, and the mean estimates are comparable with those produced by the exponential growth method. These trends are observed for each value of ρ, implying consistency in the estimates obtained with different reporting rates (Figure 2 and Supplementary Figures 4 to 6). Our adjustment does not only improve the accuracy of the central estimates of *R*_*0*_, but also improves uncertainty quantification. Average credible/confidence interval widths and coverage are reported in Figure 3. Coverage remains constant across different values of the reporting rate for the estimates obtained with the exponential growth and adjusted EpiEstim methods. On the other hand, the large bias observed in the estimates obtained with EpiEstim implies that as credible intervals get narrower, coverage decreases dramatically (Figure 3, panels B and C). The EpiEstim adjustment proposed in this paper produces generally narrower credible intervals as compared to the exponential growth method, at the price of a slight decrease in coverage. This trade-off is in avour of our adjustment for values of R_0_ larger than 2, when coverage amongst the two methods is similar. Further, we highlight that the proposed adjustment does not influence later estimates of *R*_*t*_ produced by EpiEstim (Figure 1).

**Figure 2.**
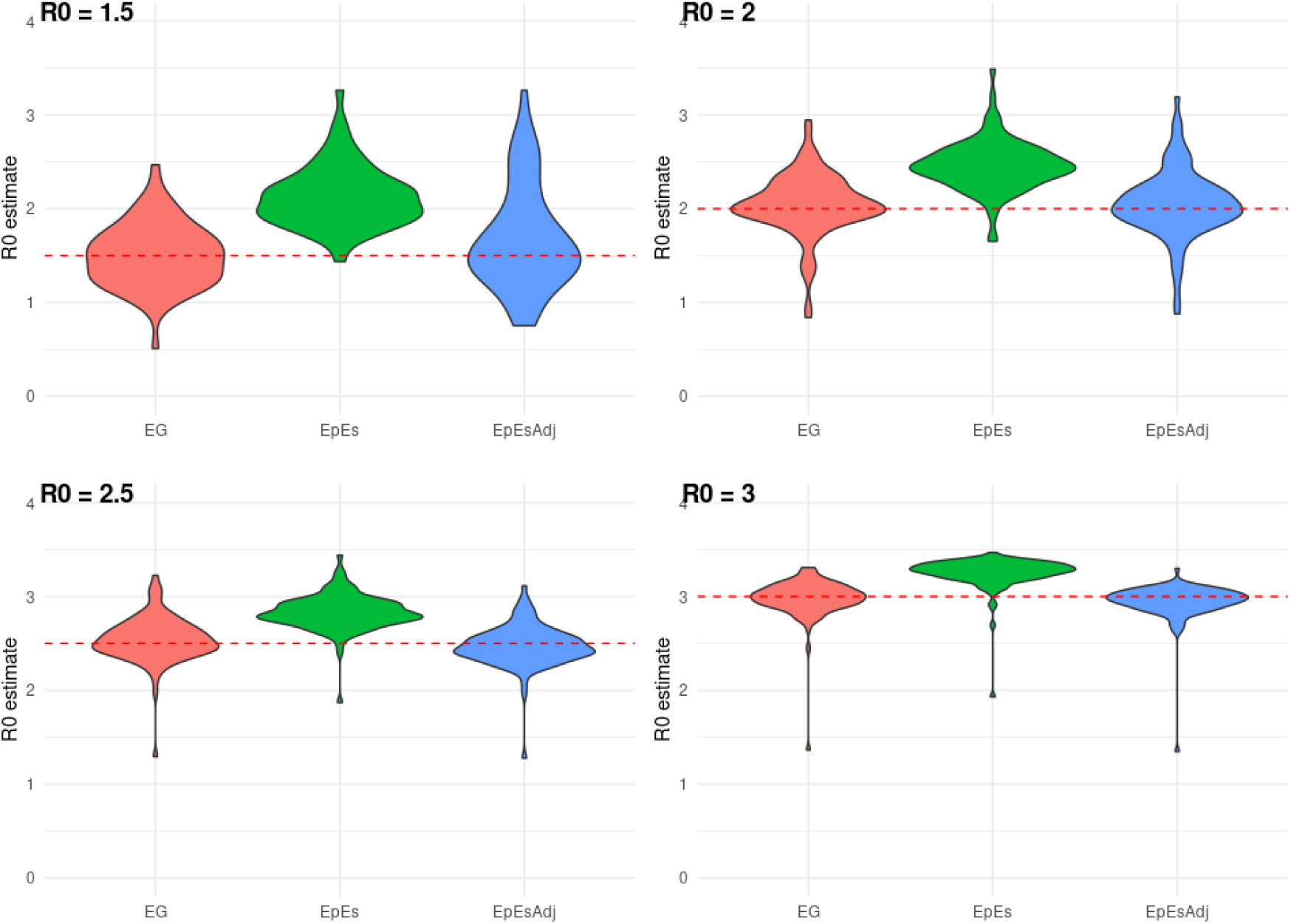
Distribution of mean R_0_ estimates assuming a fixed reporting rate ρ=50%. Each panel shows the distribution of the mean R_0_ estimates obtained using 100 simulations for a given true R_0_ value (red dashed line). Method abbreviations: Linear exponential growth rate method (EG); EpiEstim (EpEs); Adjusted EpiEstim (EpEsAdj).

**Figure 3.**
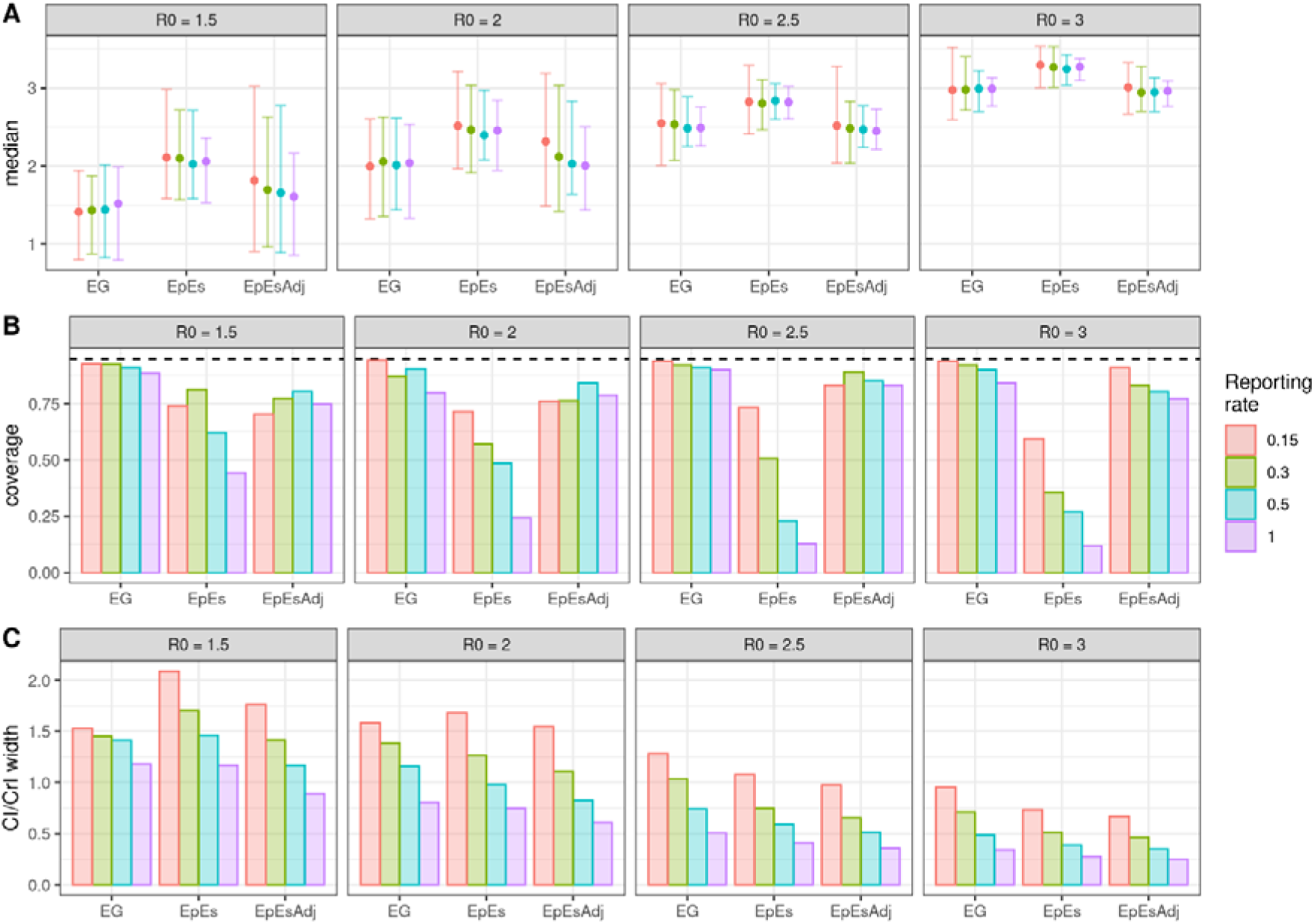
Median (point) and 95% CrI (interval range) of the basic reproduction number (R_0_) mean estimates (panel A), mean coverage (panel B) and mean 95% confidence/credible intervals widths (panel C) for different values of R_0_ and reporting rates ρ. Each panel represents distinct values of R_0_, while different colours represent different reporting rates. Method abbreviations: Linear exponential growth rate method (EG); EpiEstim (EpEs); Adjusted EpiEstim (EpEsAdj).

### Impact of missed generations

Beyond simulating undetected cases, reflecting a surveillance system which may be unprepared or unaware of a newly unfolding epidemic, we investigated the impact of the number of unobserved generations on the estimates obtained with our adjustment using simulated data generated under a scenario with perfect reporting rate (*ρ* = 1) and *R*_*0*_ = 2.5. From each epidemic trajectory, we considered 3 different left truncations of the data to account for 0, 1 and 2 unobserved generations. We then applied EpiEstim with and without the proposed adjustment using biweekly time windows starting on weeks 0, 1 and 2. Figure 4 shows the distribution of the mean estimates for the 100 simulations. Each row identifies the number of unobserved generations, while on the x-axis we show the time window used to obtain the estimate. Figure 4 also shows that the adjustment is particularly useful when larger numbers of initial generations are unobserved. While the proposed adjustment produces comparable estimates to those obtained with EpiEstim when all generations are observed (top row of Figure 4), its dependence on the exponential growth method introduces a layer of stochasticity that increases the variance of the mean estimates. On the other hand, when 1 or 2 generations are unobserved, the proposed adjustment adequately compensates for EpiEstim’s bias and the median estimates become closer to the true *R*_*0*_ value used to simulate the data.

**Figure 4.**
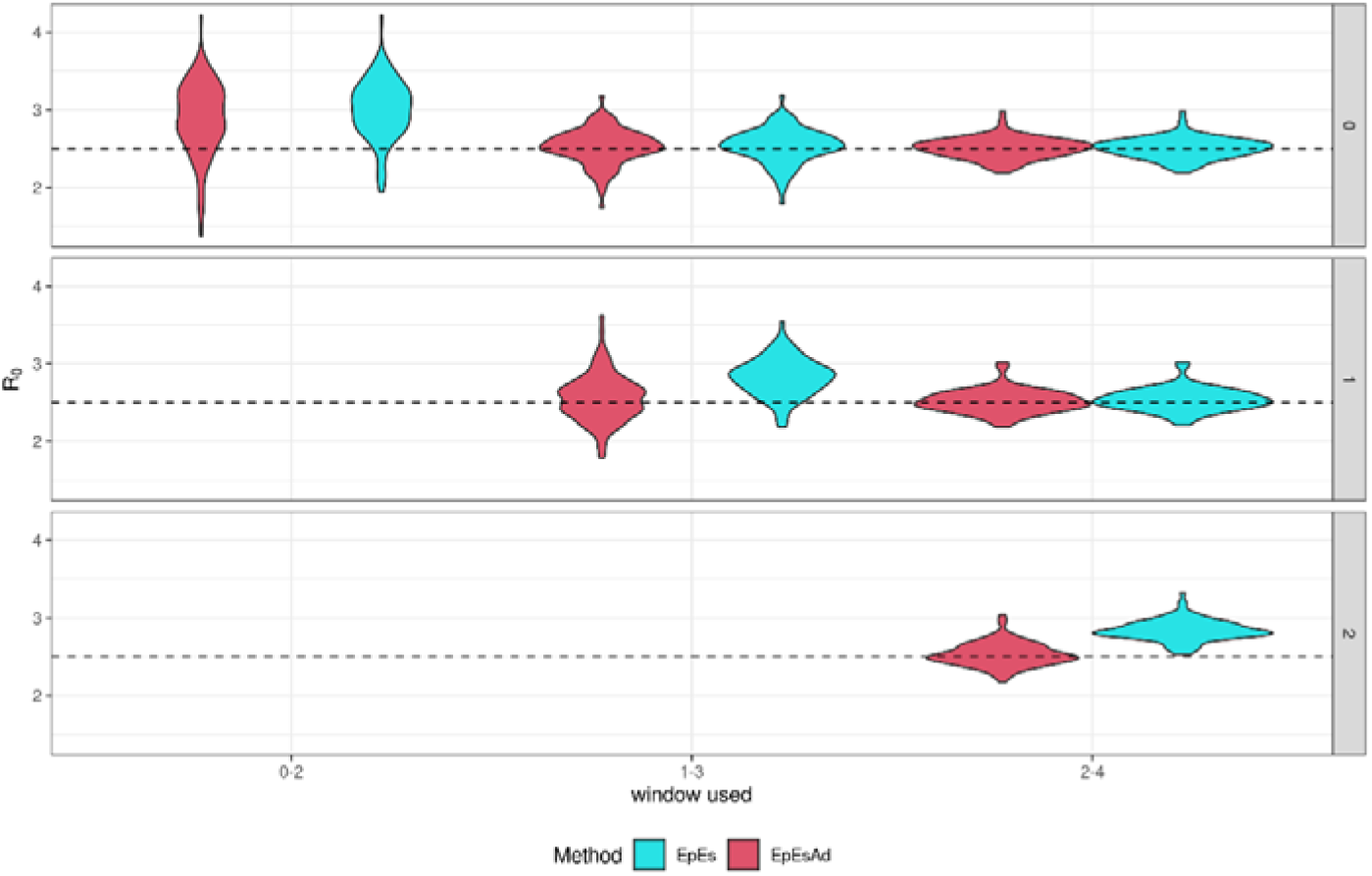
Distribution of mean R_0_ estimates, obtained using biweekly time windows (x-axis) and a variable number of unobserved generations (0, 1 and 2, see rows), assuming R_0_ = 2.5 and reporting rate ρ = 1. EpiEstim estimates are shown in blue, adjusted EpiEstim estimates are shown in red. On the axis, the sliding window used to fit the method is shown. Each window contains 14 consecutive data points, starting at week 0, or week 1, or week 2. Method abbreviations: EpiEstim (EpEs); Adjusted EpiEstim (EpEsAdj).

### Application to reported COVID-19 data

In addition to validating the proposed adjustment on simulated data, we applied it to real-world COVID-19 incidence case data reported in the John Hopkins Center for Systems Science and Engineering database [15,16]. During the initial phases of the outbreak, surveillance systems of several countries were unprepared to detect infectious individuals and it is likely that the first generations of infections were not observed, justifying our back-imputation adjustment. We selected Italy, Sweden, UK and the US as case studies. For each country, we fitted the log-transformed incidence case count reported for the first sequential 7 days of sustained transmission (no days with 0 new cases in the selected time window) with a linear regression model. We then inferred the number of unobserved cases before the selected time window and applied EpiEstim with and without adjustment to the observed and imputed data, using weekly sliding windows and assuming a generation interval with mean 5.7 days and standard deviation of 1.72 days [17]. Figure 5 shows that the adjustment lowers the estimates of *R*_*0*_ in every scenario, suggesting a role for unobserved generations of infections in overestimating the early *R*_*0*_ estimates of SARS-CoV-2, which in turn also affect the early estimates of *R*_*t*_. We obtained average *R*_*0*_ estimates of 3.6 with 95% credible interval (2.8,4.6) in the UK, 5.2 (95% CrI 4.9,5.6) for Italy, 8.7 (95% CrI 7.8,9.6) for the US and 3.9 (95% CrI 3.5, 4.3) in Sweden.

**Figure 5:**
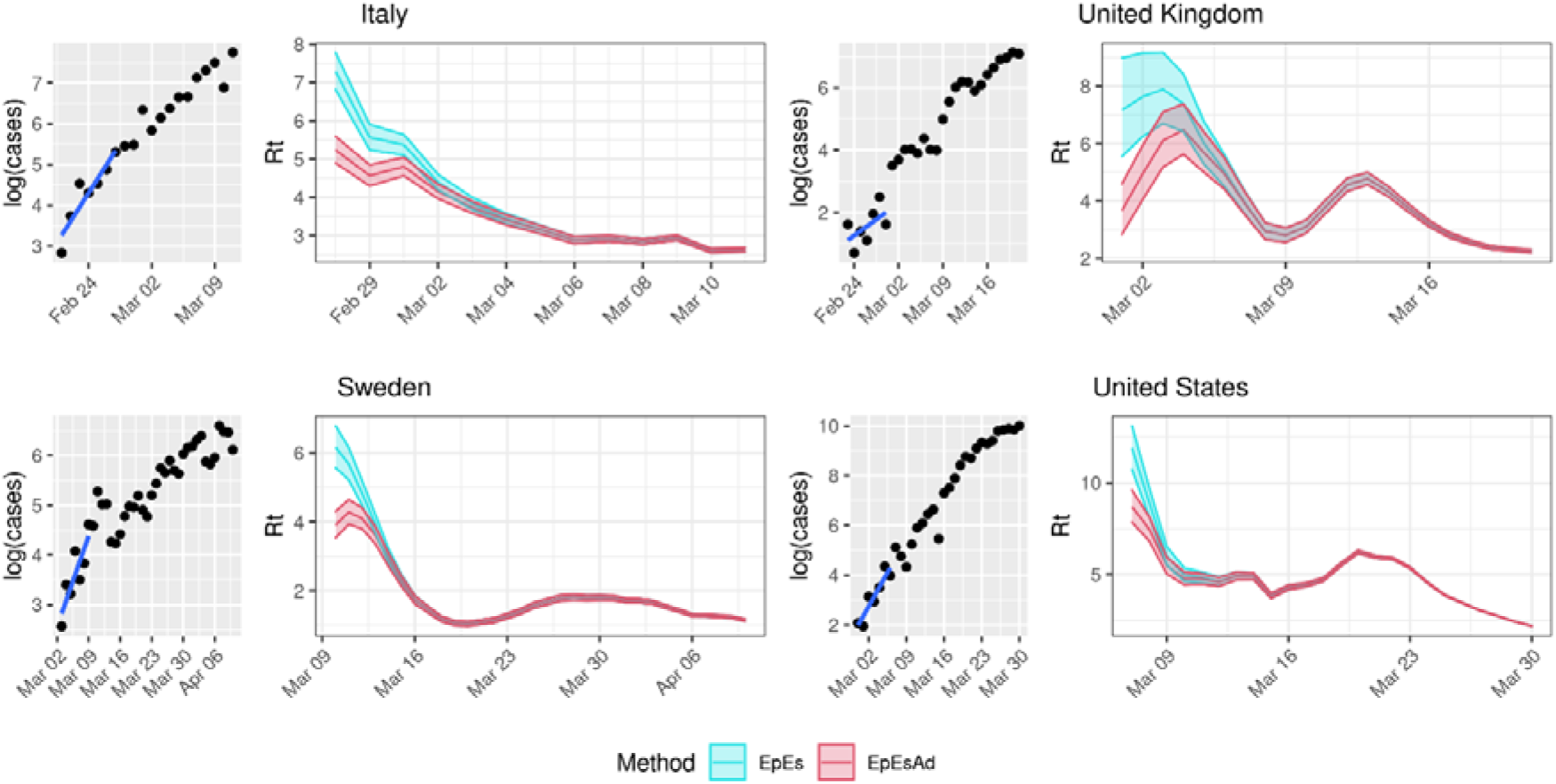
Comparison of *R*_*t*_ estimates obtained with (red) and without (blue) adjustment for COVID-19 data in 4 countries: Italy, France, the UK, and the USA. Each quadrant includes a subfigure showing the logarithm of the data and the regression line (left) and the *R*_*t*_ estimates obtained using a sliding window of 7 days (and the data up to that day) and a generation interval of mean 5.7 days and standard deviation of 1.72 days (right). Method abbreviations: EpiEstim (EpEs); Adjusted EpiEstim (EpEsAdj).

## Discussion

We propose a correction for the systematic overestimation of *R*_*0*_ that occurs in the early stages of an epidemic when using EpiEstim and other common inferential methods, utilising a back-imputation procedure which relies on exponential growth. The proposed correction aims to account for unobserved generations of infections. For this reason, we deem it to be most applicable in scenarios where generations of infections may have been missed due to emergency situations and limited testing, and generation intervals are relatively short. In practice, the adjustment will prove especially useful to evaluate transmissibility of diseases that are either asymptomatic or cause mild symptoms for a large proportion of infected individuals. Similarly, the adjustment may prove applicable to diseases characterised by long time lags between infection to symptom onset, especially if infectiousness develops significantly earlier than symptoms. In this paper, we demonstrate the application of this adjustment to the EpiEstim method, though it can be applied to other statistical methods where this bias may occur. The resulting adjusted EpiEstim method combines the best features of EpiEstim and the exponential growth method, which is more adequate for the early phases of an epidemic. In particular, the long-run *R*_*t*_ estimates of the original EpiEstim method are preserved, while the initial bias observed in the estimates is reduced by the proposed adjustment. This was confirmed in all scenarios of our simulation experiments, independently of the reproduction number and the reporting rate values used to simulate the data. The adjusted estimates of *R*_*0*_ strongly outperformed the estimates obtained with the original EpiEstim method in terms of bias and coverage, and produced tighter 95% credible intervals. Our results show that the *R*_*0*_ estimates obtained with the adjusted EpiEstim and linear exponential growth method are very similar. This is likely due, in part, to the fact that the proposed adjustment relies on linear regression, which is used for the back imputation of the unobserved generations of infections.

The proposed method does not currently incorporate uncertainty in the growth rate estimate nor in the imputed cases. Further work is required to understand how the uncertainty in our correction may be best propagated throughout the *R*_*0*_ or *R*_*t*_ estimation process.

The effect of our adjustment was evident when working with early COVID-19 data from Italy, the UK, Sweden and the US. The adjusted estimates where significantly lower than the estimates obtained with the original EpiEstim method, and were found to be largely consistent with estimates derived in Ke et al.[18]. This is especially encouraging considering that our adjusted *R*_*0*_ estimates were obtained by making use of 7 days’ worth of data, while those of Ke et al. [18] were obtained on both case and death time series data spanning months. This suggests that our method can yield improved precision with limited information, which may prove valuable in emerging epidemics. While several papers and meta-analyses reported estimates of the exponential growth rate for the 4 countries [19–21], the parameterisation of the generation interval used and the reproduction number estimates were often lacking [22] thus hindering further comparisons. Further, the adjustment alone was not enough to explain the estimated decrease in *R*_*t*_ in the study period considered (see Figure 5), possibly suggesting that changes in individual behaviour and governmental interventions lowered transmissibility of the disease. However, other biases may be playing a role, such as changes in reporting rates, overestimation due to importations [23], or model misspecification.

Critically, the results are sensitive to the choice of EpiEstim parameters, such as the generation interval and the length of the time window used. As expected, longer generation intervals produce larger reproduction number estimates, and we observe larger discrepancies between the estimates obtained with the original and adjusted EpiEstim methods, even if the coefficient of variation is kept constant (Supplementary Figures S1-S3).

Concerning the choice of the sliding window, the larger time windows produce smoother estimates, which reduces the impact of the imputed cases. This means that our adjustment has a much smaller effect when considering sliding windows covering two generation intervals worth of data or more.

It is possible that other methods may estimate the initial growth rate more accurately than linear regression particularly if the assumption of exponential growth is not met. In fact, the linear regression model assumes normality of the errors, which may not always hold true with count data. In future work it will be interesting to compare alternative methods to back impute unobserved cases from early incidence data.

Here, we have used a linear exponential growth method to infer missing initial generations of infection, improving the accuracy of early *R*_*0*_ and *R*_*t*_ estimates produced by commonly used statistical methods such as EpiEstim. Our analysis shows how a simple adjustment can reduce initial bias in reproduction number estimates in a newly emerging epidemic when the interpretation of reproduction number estimates is crucial for public health decision making.

## Data Availability

All data used and produced in the present work are available at: https://github.com/abriz97/R0_simulations

https://github.com/CSSEGISandData/COVID-19

## Contributions

AB, MO’D, ID conceived the study and the methods; AB performed the analysis, developed the code and wrote the first draft; all authors revised and approved the manuscript.

## Data Statement

The data and R code to reproduce this analysis are available on GitHub: https://github.com/abriz97/R0_simulations

