## Supplementary Material for "Refining reproduction number estimates to account for unobserved generations of infection in emerging epidemics"

### **Supplementary Information: Refining reproduction number estimates to account for unobserved generations of infections in emerging epidemics**

Authors: Andrea Brizzi^1+^, Megan O’Driscoll^2,3+^, Ilaria Dorigatti^2^

Author Affiliations:
^1^Department of Mathematics, Imperial College London, London, United Kingdom

^2^MRC Centre for Global Infectious Disease Analysis and Jameel Institute, School of Public Health, Imperial College London, London, United Kingdom
^3^Department of Genetics, University of Cambridge, Cambridge, United Kingdom

^+^ Equal contribution

*Sensitivity analysis on the generation interval distribution*

We conducted a sensitivity analyses to assess the impact of the assumed length of the mean generation interval on the reproduction number estimates obtained with EpiEstim and our proposed adjustment. We used the COVID-19 data described in the main section and considered three different possible mean distributions with the same coefficient of variation set to 2. In Figure S1 the generation interval mean and standard deviation are set to 4 and 2 respectively, in Figure S2 the generation interval mean and standard deviation are set to 6 and 3 respectively and Figure S3 the generation interval mean and standard deviation are set to 8 and 4 respectively.

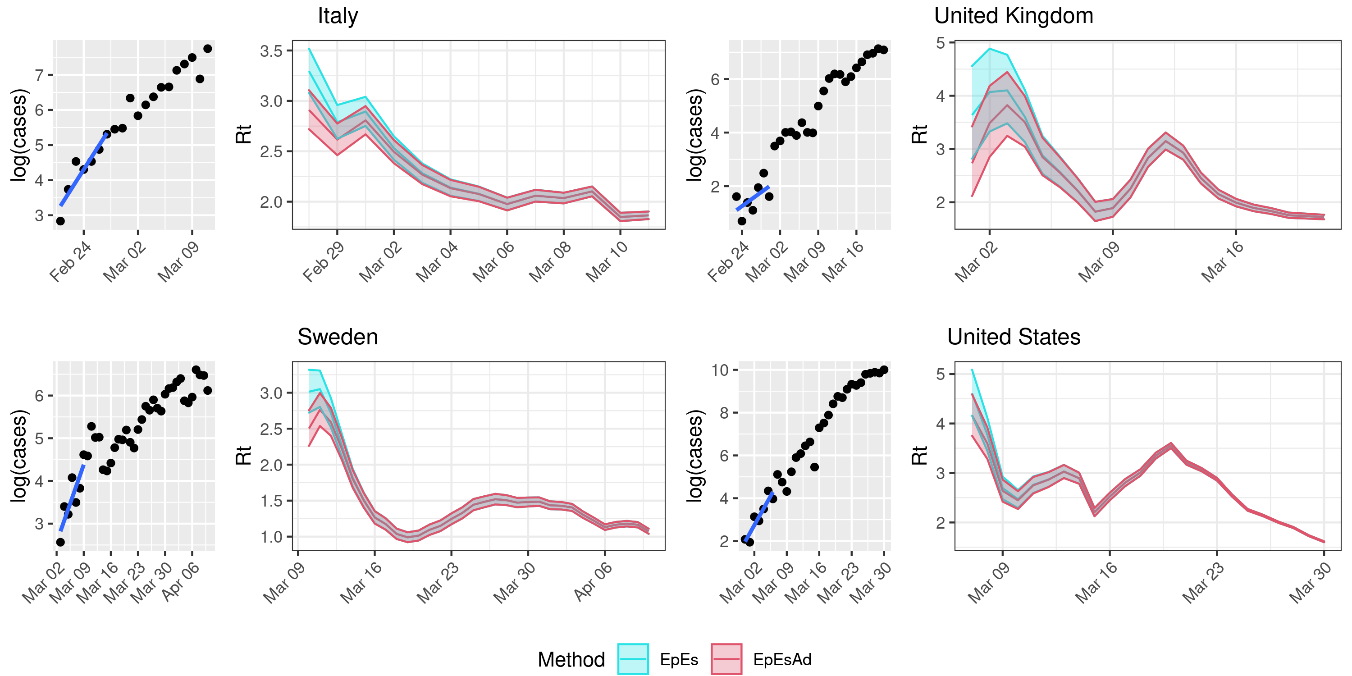

**Figure S1. Sensitivity analysis on generation interval**. Each quadrant includes a subfigure showing the logarithm of the data and the regression line (left) and the *R_t_* estimates obtained using a sliding window of 7 days (and the data up to that day) and a generation interval of mean 4 days and standard deviation of 2 days (right). Method abbreviations: EpiEstim (EpEs); Adjusted EpiEstim (EpEsAdj).

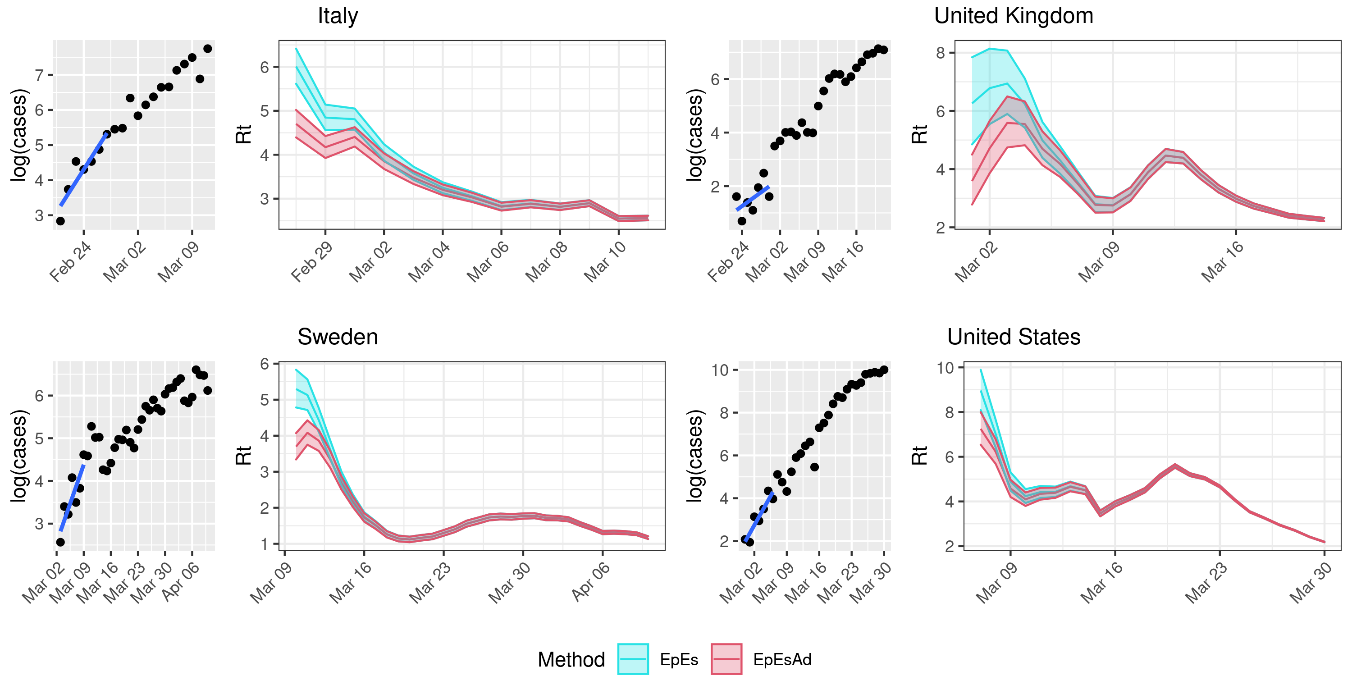
**Figure S2. Sensitivity analysis on generation interval.** Each quadrant includes a subfigure showing the logarithm of the data and the regression line (left) and the *R_t_* estimates obtained using a sliding window of 7 days (and the data up to that day) and a generation interval of mean 6 days and standard deviation of 3 days (right). Method abbreviations: EpiEstim (EpEs); Adjusted EpiEstim (EpEsAdj).

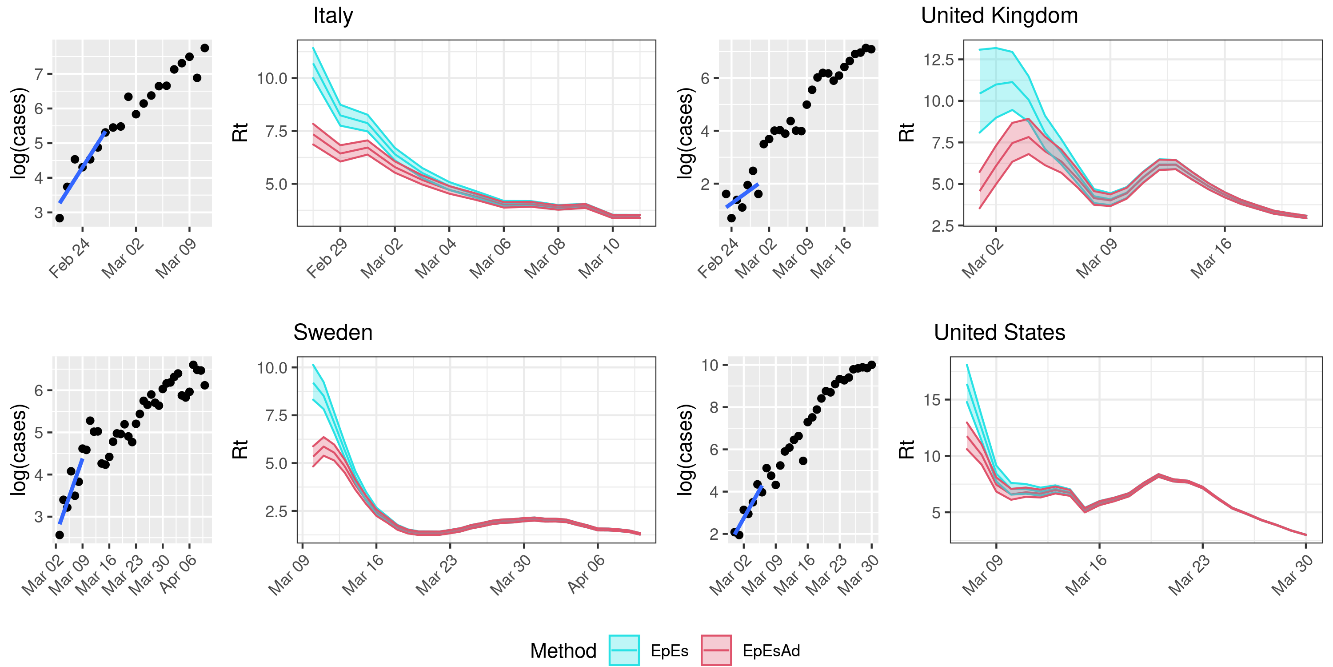

**Figure S3. Sensitivity analysis on generation interval.** Each quadrant includes a subfigure showing the logarithm of the data and the regression line (left) and the *R_t_* estimates obtained using a sliding window of 7 days (and the data up to that day) and a generation interval of mean 8 days and standard deviation of 4 days (right). Method abbreviations: EpiEstim (EpEs); Adjusted EpiEstim (EpEsAdj).

*Sensitivity analysis on the assumed reporting rate*

We also performed a sensitivity analysis to assess the impact of the assumed reporting rate on the *R_0_* estimates obtained in our simulations. The results are shown in Figures S4-S6.

Figure S4 considers a reporting rate set to 15%, Figure S5 considers a reporting rate set to 30%, while Figure S6 considers a reporting rate of 100%.

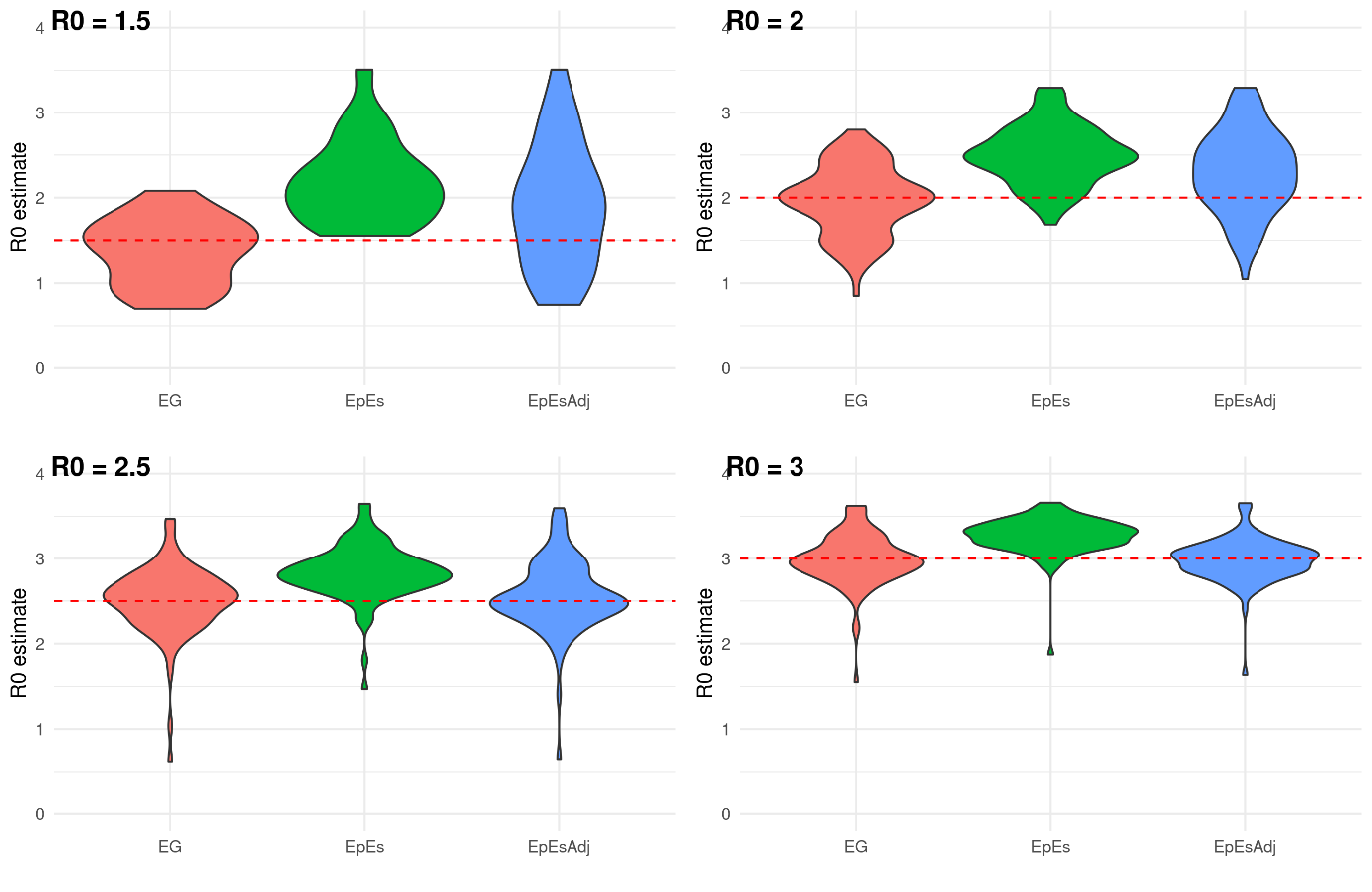
**Figure S4. Sensitivity analysis on reporting rates.** Distribution of mean R_0_ estimates assuming a fixed reporting rate ρ=15%. Each panel shows the distribution of the mean R_0_ estimates obtained using 100 simulations for a given true R_0_ value (red dashed line). Method abbreviations: Linear exponential growth rate method (EG); EpiEstim (EpEs); Adjusted EpiEstim (EpEsAdj).

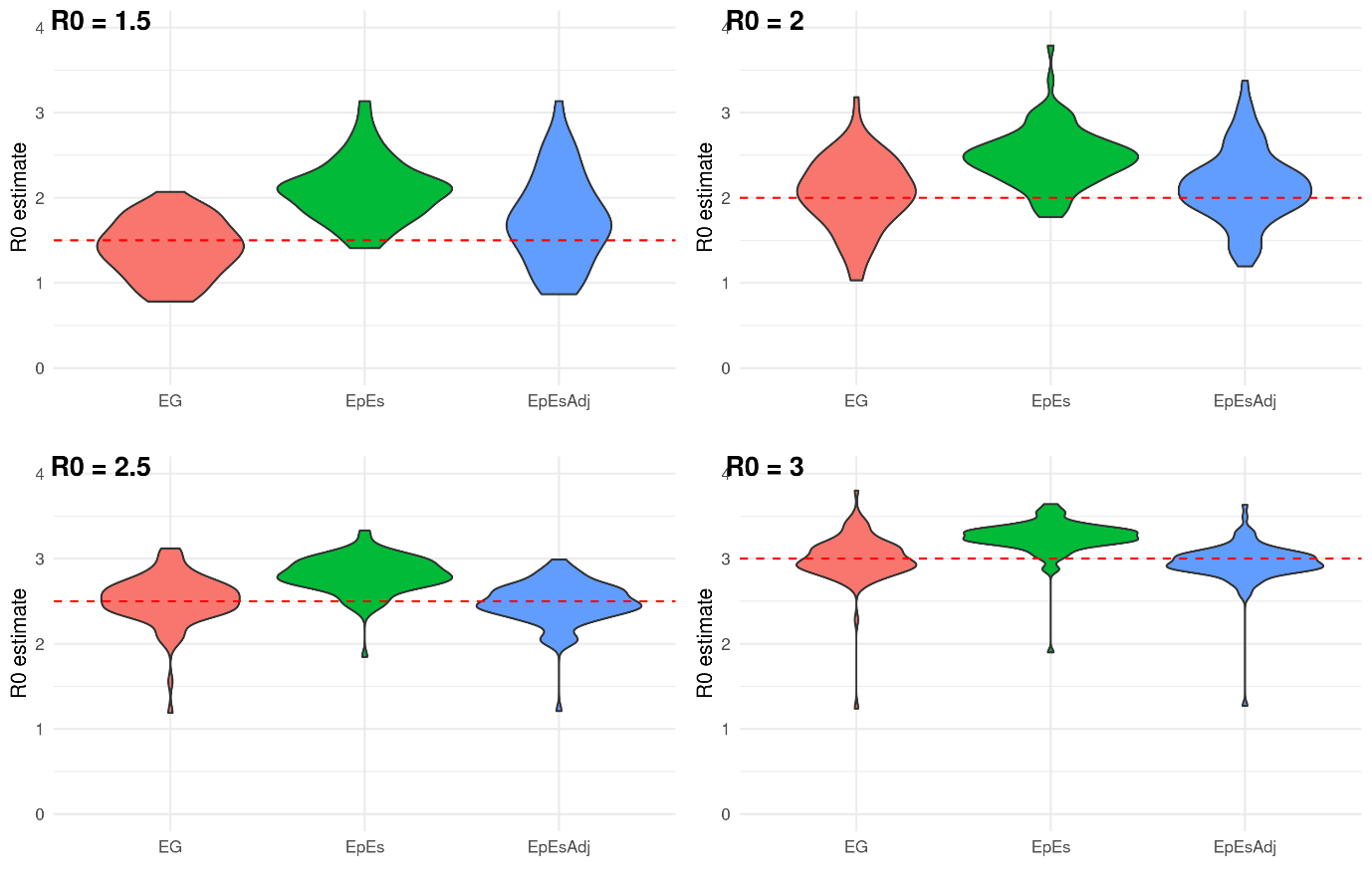
**Figure S5. Sensitivity analysis on reporting rates.** Distribution of mean R_0_ estimates assuming a fixed reporting rate ρ=30%. Each panel shows the distribution of the mean R_0_ estimates obtained using 100 simulations for a given true R_0_ value (red dashed line). Method abbreviations: Linear exponential growth rate method (EG); EpiEstim (EpEs); Adjusted EpiEstim (EpEsAdj).

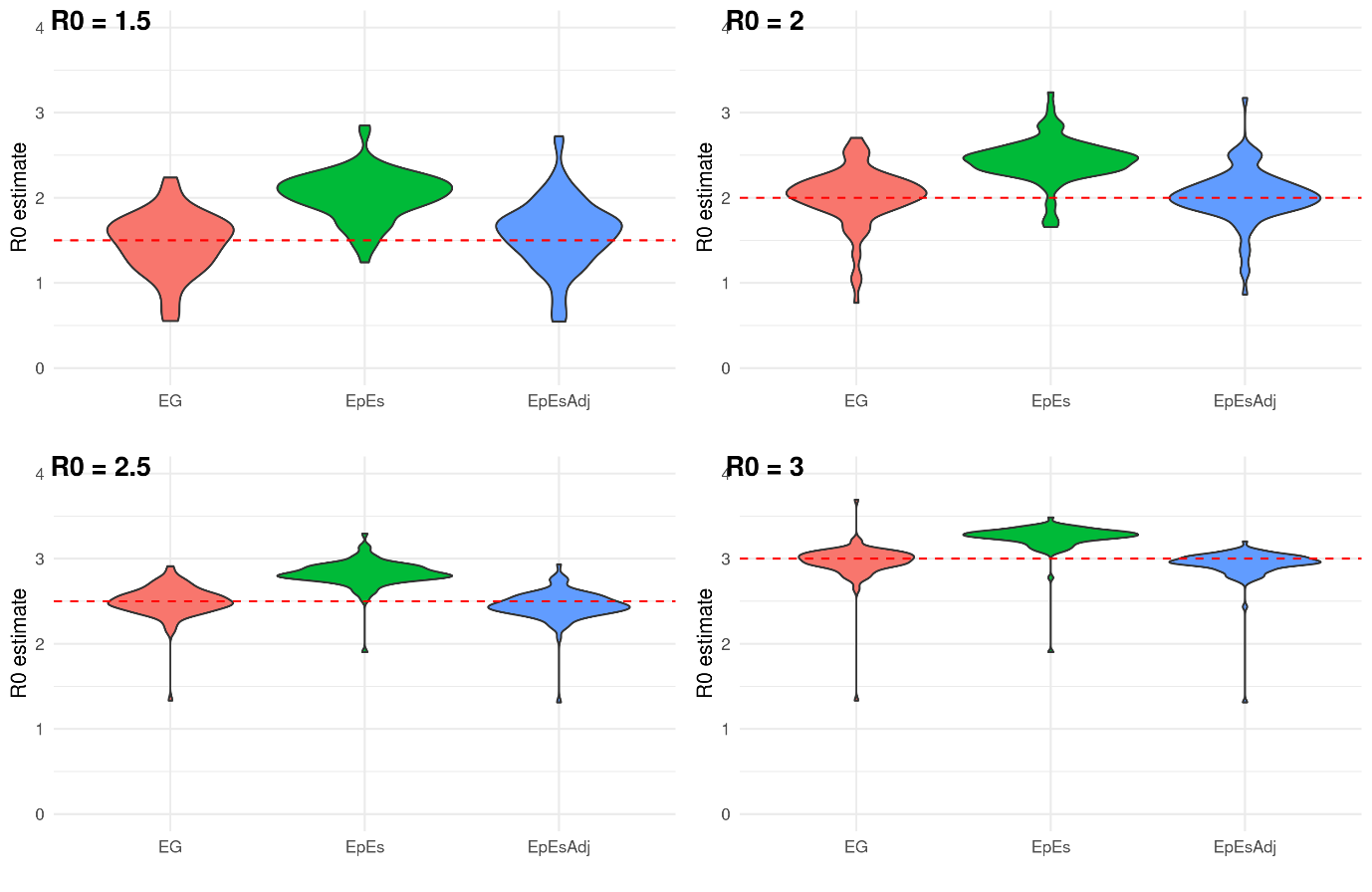
**Figure S6. Sensitivity analysis on reporting rates.** Distribution of mean R_0_ estimates assuming a fixed reporting rate ρ=100%. Each panel shows the distribution of the mean R_0_ estimates obtained using 100 simulations for a given true R_0_ value (red dashed line). Method abbreviations: Linear exponential growth rate method (EG); EpiEstim (EpEs); Adjusted EpiEstim (EpEsAdj).
